# Characterization of oscillations in the brain and cerebrospinal fluid using ultra-high field magnetic resonance imaging

**DOI:** 10.1101/2023.12.05.23299452

**Authors:** Tiago Martins, Tales Santini, Bruno de Almeida, Minjie Wu, Kristine A. Wilckens, Davneet Minhas, James W. Ibinson, Howard J. Aizenstein, Tamer S. Ibrahim

## Abstract

Development of innovative non-invasive neuroimaging methods and biomarkers are critical for studying brain disease. In this work, we have developed a methodology to characterize the frequency responses and spatial localization of oscillations and movements of cerebrospinal fluid (CSF) flow in the human brain. Using 7 Tesla human MRI and ultrafast echo-planar imaging (EPI), *in-vivo* images were obtained to capture CSF oscillations and movements. Physiological data was simultaneously collected and correlated with the 7T MR data. The primary components of CSF oscillations were identified using spectral analysis (with frequency bands identified around 0.3Hz, 1.2Hz and 2.4Hz) and were mapped spatially and temporally onto the MR image domain and temporally onto the physiological domain. The developed methodology shows a good consistency and repeatability (standard deviation of 0.052 and 0.078 for 0.3Hz and 1.2Hz bands respectively) *in-vivo* for potential brain dynamics and CSF flow and clearance studies.

## 1. Introduction

Clearance and exchange of brain fluids promote brain health by removing neurotoxic metabolic byproducts from the brain such as amyloid beta and tau (Nedergaard, 2013; Xie et al., 2013). Clearance of brain fluids is driven by pulsations of the perivascular space from autonomic nervous system (ANS) activity, which may vary as a function of brain states such as sleep and wakefulness (Hauglund et al., 2020; Herculano-Houzel, 2013; Xie et al., 2013) as well as brain diseases such as Alzheimer’s disease (Peng et al., 2016; Ramanathan et al., 2015) and major depressive disorder(Hock et al., 1998; Pomara et al., 2012). The lymphatic draining system of the brain tissue, also known as the glymphatic system, is described as convection of cerebrospinal fluid (CSF) between the peri-arterial and peri-venous spaces. This convective flow is partially due to the cardiac-induced blood flow pulsations along the arteries (Adolph et al., 1967; Iliff et al., 2013; Martin et al., 2012; Schroth & Klose, 1992). Water is propelled by the arterial pulsations through aquaporin channels and supports solute transport from extracellular interstitial spaces, through perivascular spaces, and into CSF spaces. CSF and waste products from the brain are then pushed from parenchyma to subarachnoid spaces and eventually cleared via arachnoid granulations and dural and nasal lymphatic vessels (Iliff et al., 2013; Kiviniemi et al., 2016; Leon et al., 2017; Rennels et al., 1990).

Development of quantitative CSF imaging methods is critical to understand factors that influence brain fluid clearance. T1-weighted magnetic resonance imaging (MRI) with intrathecal injection of a gadolinium (Gd)-based contrast agent has been used to characterize CSF flow in human participants with idiopathic normal pressure hydrocephalus (iNPH) and dementia (Eide & Ringstad, 2019; Ringstad et al., 2017, 2018). This technique has afforded fully quantitative, high-resolution imaging of CSF and interstitial fluid (ISF) flow throughout the whole head but is highly invasive due to necessary lumbar puncture. Gd may also deposit in the brain, inhibiting longitudinal study (Gulani et al., 2017).

Fast acquisition functional magnetic resonance imaging (fMRI) paradigms have also been used to characterize CSF dynamics in iNPH and Alzheimer’s disease (AD) patients and during sleep (Fultz et al., 2019; Shanks et al., 2019; Yamada et al., 2020; Yang et al., 2022). While non-invasive, these sequences have relatively poor signal-to-noise ratio and spatial resolution and have been limited to narrow fields of view encompassing only the 4^th^ ventricular or cerebral aqueduct.

The bulk changes in blood volume at the capillary level could cause widespread fluctuations of measured signal intensity with the cardiac cycle. Furthermore, large vessel pulsatility may cause tissue movement and production of an influx of unsaturated blood into the slice of interest affecting the measured signal intensity in the areas adjacent to the vessels. This leads to a signal variation when using T2* echo-planar imaging (EPI) acquisitions (Dagli et al., 1999). Hence, fMRI and other techniques have been used to characterize different sources of pulsations in the brain (Biswal et al., 1995; Dagli et al., 1999; Kiviniemi et al., 2000; Poncelet et al., 1992; Purdon & Weisskoff, 1998). Thus, using MRI of CSF dynamics can highly inform the study of brain diseases and the role of sleep-wake states (Fultz et al., 2019; Xie et al., 2013).

Ultra-high field MRI (≥ 7 Tesla) provides a major advantage of increased signal-to-noise ratio (SNR). The enhanced SNR can be used either to increase the resolution of the images or to decrease the scanning time (with the use of higher acceleration factors). Other advantages of 7 Tesla (T) field strength are the higher sensitivity to blood-oxygen-level-dependent (BOLD) signal and better vasculature conspicuity (Moser et al., 2012; Santini, Wood, et al., 2021). The high signal-to-noise ratio (SNR) and fast acquisitions of 7T MRI scanners allow studies to perform analysis of blood and cerebrospinal fluid (CSF) flow within the brain (Scouten & Constable, 2008).

This work revolves around the creation of a method of acquisition and analysis that can be used as biomarker for study of central nervous system functioning and brain diseases. Using fast EPI and physiological acquisitions, we viewed and analyzed the CSF MR signal in real-time. We report CSF oscillation patterns through spectral analysis and apply the same methodology across different datasets to validate the observed results.

## 2. Methods

The overall design of this study is based on two main steps: in-vivo 7T image acquisition, concurrent with physiology measurements, and image processing with spectral analysis. The processing and analysis of the power-frequency spectrum and its corresponding spatial mappings were fine tuned for the processing parameters such as detection bandwidth and peak span, thresholding levels for masks and smoothing degrees for filtering.

The volunteers scanned for this work provided informed consent as part of an approved study by the University of Pittsburgh’s Institutional Review Board (identification number PRO17030036). For CSF flow data collection, five healthy volunteers (all females, age range 21-25 years old) were scanned to obtain EPI data from the whole brain, including the cerebellum. For comparison between physiological and MRI data, one additional healthy volunteer (male, age range 26-30 years old) was scanned and had physiological data collected simultaneously.

### 2.1 Image Acquisition

Images were acquired using a whole-body 7T MRI system (Siemens 7T MAGNETON) and with the human-connectome EPI multiband MR sequence (Moeller et al., 2010; Uğurbil et al., 2013). The sequence is capable of achieving fast acquisitions, high contrast to the CSF flow, and high sensitivity to BOLD signal, thus making it a good candidate for studies of sleep and neurodegenerative and psychological disorders. The imaging was done with an in-house developed and fabricated 16-channel Tic-Tac-Toe transmit array with a 32-channel receive head coil (Krishnamurthy et al., 2019; Santini et al., 2018; Santini, Wood, et al., 2021) that is load insensitive (Ibrahim et al., 2008; Kim et al., 2016; Santini, Wood, et al., 2021) and capable of whole brain homogenous imaging at 7T (Ibrahim et al., 2013). By using this coil design, we were able to acquire signal from the entire brain with minimal regions of significant excitation losses using the single transmit mode of the 7T scanner.

The acquired EPI images yield a real-time visualization of the CSF flow in the brain. The sequence was optimized to perform fast brain imaging. The main data acquisition was done with echo time (TE) of 20 ms (carefully chosen for future potential BOLD analysis), repetition time (TR) of 155ms, isotropic resolution of 2mm, and acceleration factor of 2. The field of view (FOV) was 192 mm x 192 mm x 6 mm. The acquisition was broken into 19 slabs of 3 axial slices each for a total of 57 slices, which is enough to have whole-brain coverage. A total of 600 volumes were sequentially acquired per slab in a single sequence run for an acquisition time of 1 minute and 36 seconds per slab.

During the development of the protocol, the EPI acquisition on the Volunteer 1 was done using TR of 152ms but only 15 slabs for a total of 45 axial slices. Another EPI acquisition on the same volunteer was also performed using TR of 51ms and a single slice.

Two spin-echo echo-planar images were also acquired for B_0_ field distortion correction with the same phase encoding (PE) direction of the EPI acquisition (PA) and with the opposite PE direction (AP). These acquisitions were performed for 57 slices, TE of 39.4ms, TR of 6000ms with matched parameters as the EPI sequence in terms of field of view, resolution, number of slices, echo spacing, and position.

A T1-weighted imaging (MPRAGE) sequence was used for proper localization of the EPI field of view and as a structural scan for the image processing. This acquisition was done using 0.75mm isotropic resolution, TR of 3000ms, TE of 2.17ms, and 256 slices for a coverage of 240 mm x 173 mm x 192 mm in total time of acquisition of ∼5 minutes.

### 2.2 Physiological Measurements

Electrocardiogram (ECG) and respiration signals were collected from a consented volunteer inside the MR scanner using MR compatible ECG leads and an expansion belt attached to the chest to track inflation and deflation of the chest during respiration. Acquisition was digitalized using BIOPAC system (“ECG,” n.d.; “Respiration Transducer for MRI | TSD221-MRI | Research | BIOPAC,” n.d.). The simultaneously collected data allows temporal signal analysis of both MR and physiologic signals. The imaging data acquired in conjunction with the physiological data used TR of 75ms, TE of 28ms, and slice thickness of 4 mm.

### 2.3 Image Processing

The processing pipeline was developed based on MATLAB (*MATLAB - MathWorks*, n.d.), ANTs (Avants et al., 2009), and FSL (Jenkinson et al., 2012) software. It consists of denoising, distortion correction, bias correction, and skull stripping of each dataset volume. The initial step was loading the slabs and merging them into a single dataset. Next, denoising was performed using a noise estimation tool with variance stabilization transformation (VST) for Rician-distributed noise (Foi, 2011). The Rician heteroscedastic noise is converted to a homoscedastic noise after the forward VST. The block-matching 4D (BM4D) denoising algorithm (Maggioni et al., 2013) can then be applied and the denoised image is obtained after the inverse VST. This tool has been used for other MRI applications(Santini, Koo, et al., 2021) and yields a good result when applied to EPI data. Distortion correction was done using the estimated B_0_ maps derived from the spin-echo sequence using *topup* tool (Andersson et al., 2003) from FSL software package. The generated map was used for correction of the EPI data. Then, the images were bias corrected using the N4(Tustison et al., 2010) tool from the ANTs software package with spline distance parameter of 200. The final skull stripping was performed using the FSL brain extraction tool (BET).

### 2.4 Spectral Analysis

The frequency analysis was performed for each dataset individually and resulted in both a frequency power spectrum and a mask for brain localization of specific frequency bands. A frequency spectrum was calculated for selected points for validation of the findings across different brain regions.

After processing each individual slab of the EPI data, the frequency processing and analysis were performed in MATLAB and Python. The time series of each voxel was used to generate a frequency spectrum using fast Fourier transform (FFT). With the 600 volumes of 155ms TR acquisition, the frequency resolution of the frequency spectrum is 0.011Hz and the maximum frequency is 3.23Hz. The same frequency analysis for the 51ms TR data produces a much larger frequency spectrum of up to 9.8Hz. Therefore, frequency components higher than 3Hz can be observed and analyzed. The analysis was done using both the average of the 3D space and individual points manually selected.

Spatial analysis was done by creating image masks based on the localization of voxels with significant signal in each frequency band. Each frequency band was defined with a bandwidth of 0.3Hz and was centered at the local maxima of the frequency spectrum amplitude with a minimum peak distance 0.15Hz (0.5Hz was used for the dataset with 51ms TR). The power map of a given frequency band was determined voxel-wise by averaging respective power values within the frequency band. For better visualization, each power map was then binarized with a chosen threshold (75% of the peak amplitude of the corresponding frequency band) and spatially smoothed using a Gaussian filter (sigma of 1.6), generating the final masks for each frequency band. These masks were overlaid on the original EPI and T1 weighted acquisitions for anatomical reference. The T1 weighted image was registered with the average EPI image of the dataset using SPM12.

## 3. Results

A video was created based on the image series of the fast EPI data after acquisition and processing (Figure 1). The video visually indicates the presence of periodical signal from the CSF flow. To confirm the presence of physiological signals such as respiration and cardiac motion, CSF temporal data was aligned with measurements from the electrocardiogram and respiration belt for visual comparison of similarity between the physiological activities and the change in signal intensity from CSF regions (Figure 2a). The frequency spectrum of the datasets was also aligned following the same comparison as the time series data (Figure 2b). The two major signal bands were highlighted between the ECG and CSF data (around 1.1Hz) and the respiration belt and CSF data (around 0.3Hz).

**Figure 1:**
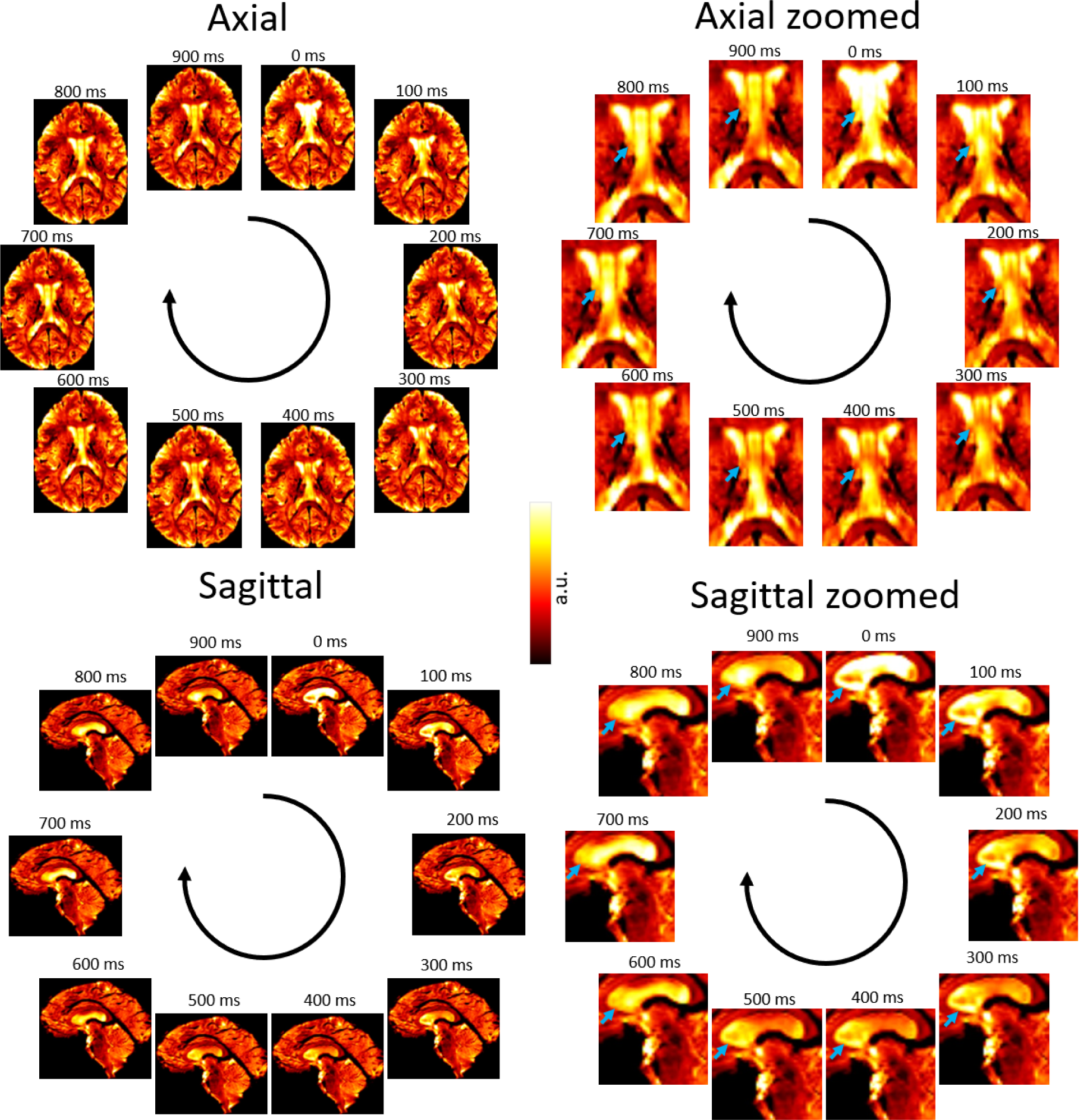
Fast EPI acquisition (TR=100ms) showing signal changes due to CSF flow; axial slices with a spatial resolution of 1.53 x 1.53 x 3mm and a sagittal slice with spatial resolution of 1.5 x 1.5 x 4.4mm. The blue arrows point to regions of large variation in signal over time. A video showing these oscillations in real time is available at https://doi.org/10.6084/m9.figshare.24022932

**Figure 2:**
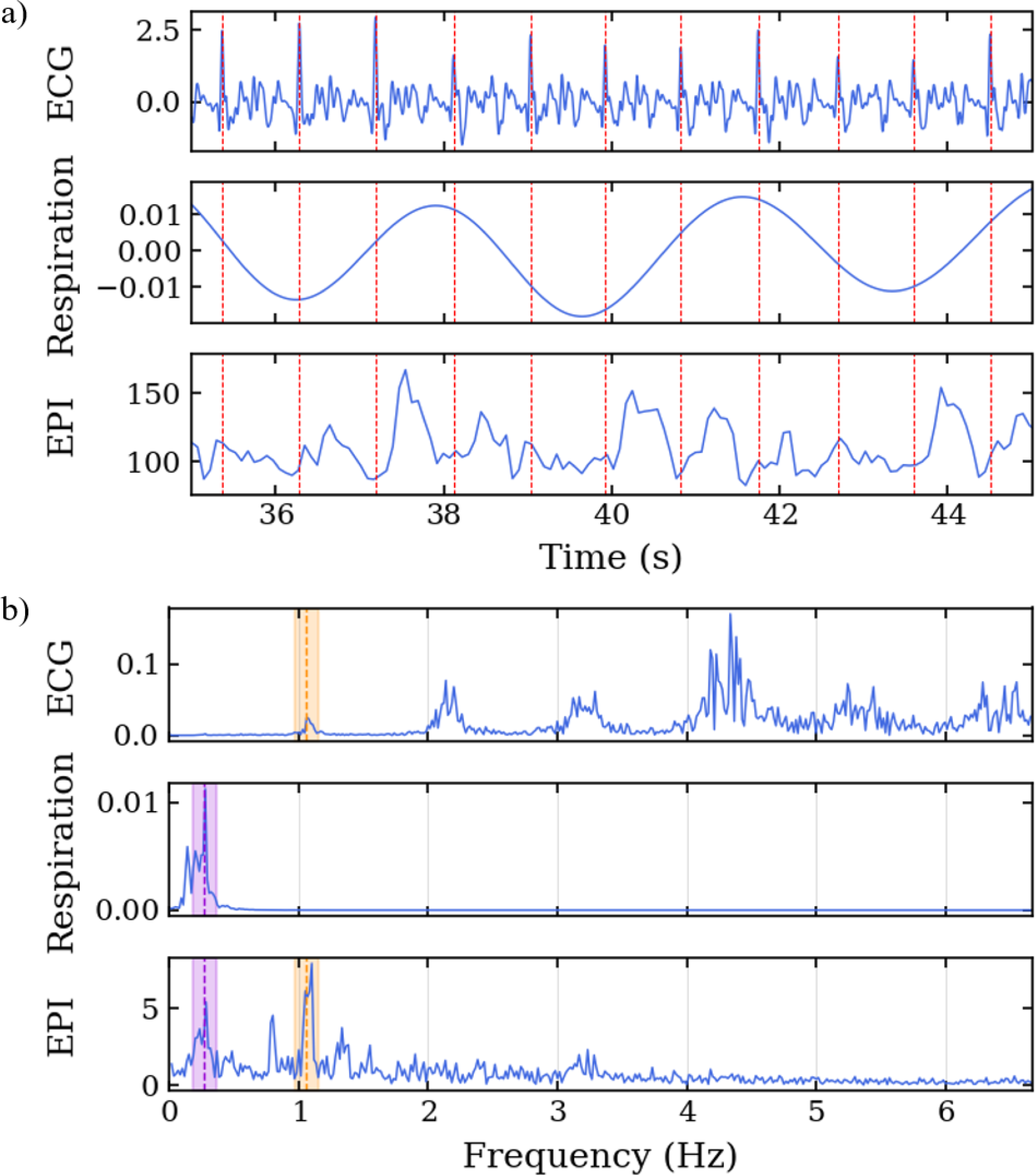
Data acquisition along the cardiac cycle. Echo-planar imaging acquisition performed with concurrent physiological measurement of electrocardiogram in the 7T scanner. a) Time series of ECG, respiration belt, and EPI signals. The EPI signal was temporally aligned with the physiological data using an external trigger signal from the scanner. The red lines represent the R-peaks of the ECG. B) Frequency spectrum of the ECG, respiration belt, and EPI signals. The purple region highlights the common frequency band between the respiration and EPI signals, and the orange region highlights the common frequency band between the ECG and EPI signals.

To verify that various points of the brain contribute differently on the frequency spectrum, Figure 3 represents the frequency spectrum for 9 arbitrary points throughout the brain. The position of each point is described by the brain anatomy that it belongs to as shown on the top-right corner of each spectrum graph. Most of the points show frequencies around 1.2Hz. Depending on the position, the signal shows the 0.3Hz and/or the 2.4Hz bands.

**Figure 3:**
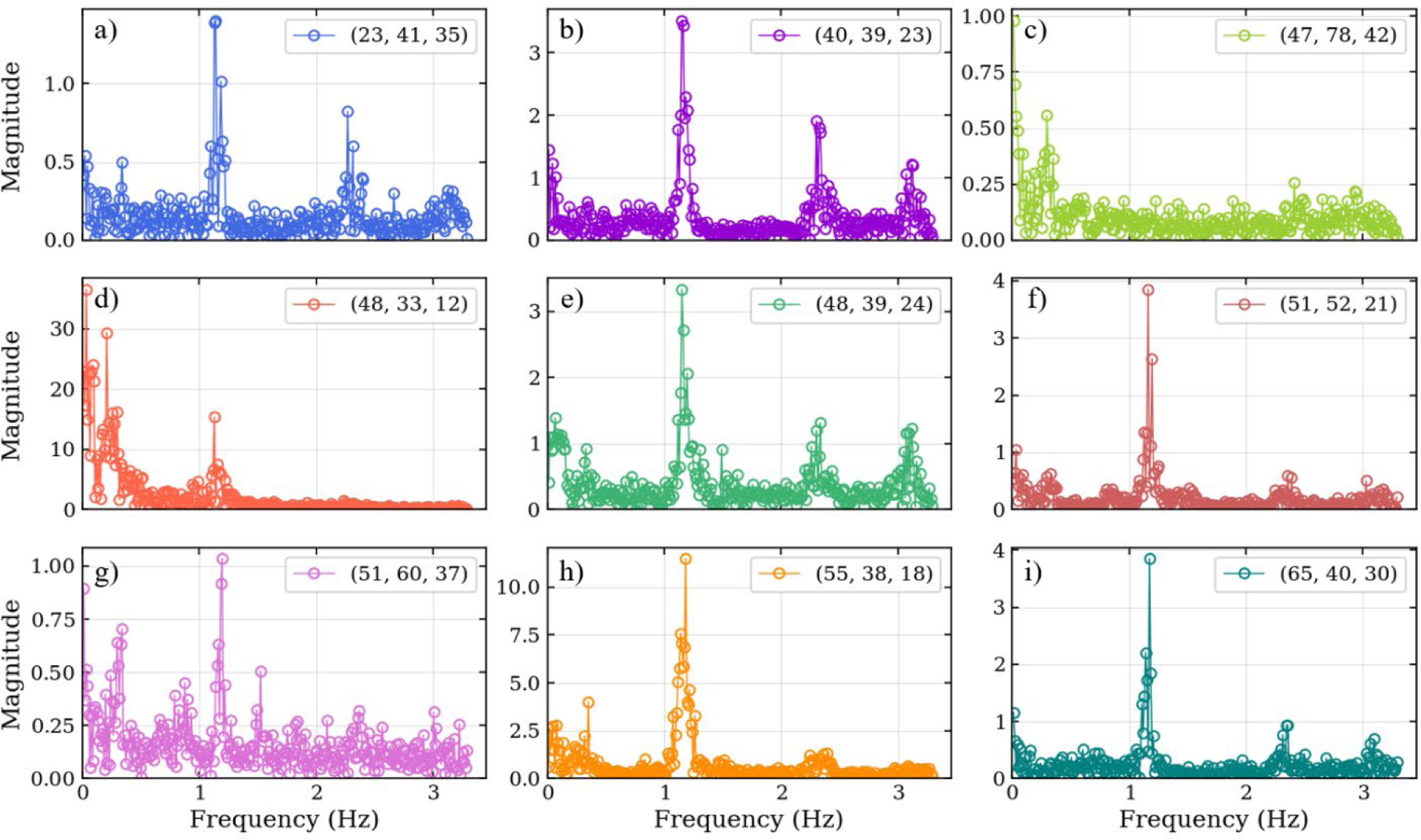
Frequency spectrum for 9 selected points (a-i) throughout the brain for volunteer 1. Some points show higher intensity on the 1.2Hz and 2.4Hz bands (points a, b, e, f, g, h, and i) whereas other show more on the 0.3Hz band (points c, d, and g). The labels describe the brain anatomy where the data was obtained.

A frequency analysis was obtained from the EPI data. For each participant, the frequency spectrum was calculated for the average of all voxels in the dataset to validate the observations (Figure 4). The most significant frequency bands were highlighted after identification of the center frequency using a local maxima approach (*findpeaks* implementation in Python). For all volunteers, bands with similar center frequencies of approximately 0.3Hz, 1.2Hz, and 2.4Hz can be identified. Table 1 shows the center frequency for the bands calculated for each volunteer. The frequency bands with centers at 0.3Hz and 1.2Hz closely approximate the respiration and cardiac frequencies of a human adult (around 18 breaths per minute and 72 heart beats per minute, respectively). These bands can be identified as Band 1 and Band 3 on Table 1. The averages for all volunteers are 0.322Hz and 1.217Hz respectively with a standard deviation of 0.052 and 0.078 respectively.

**Figure 4:**
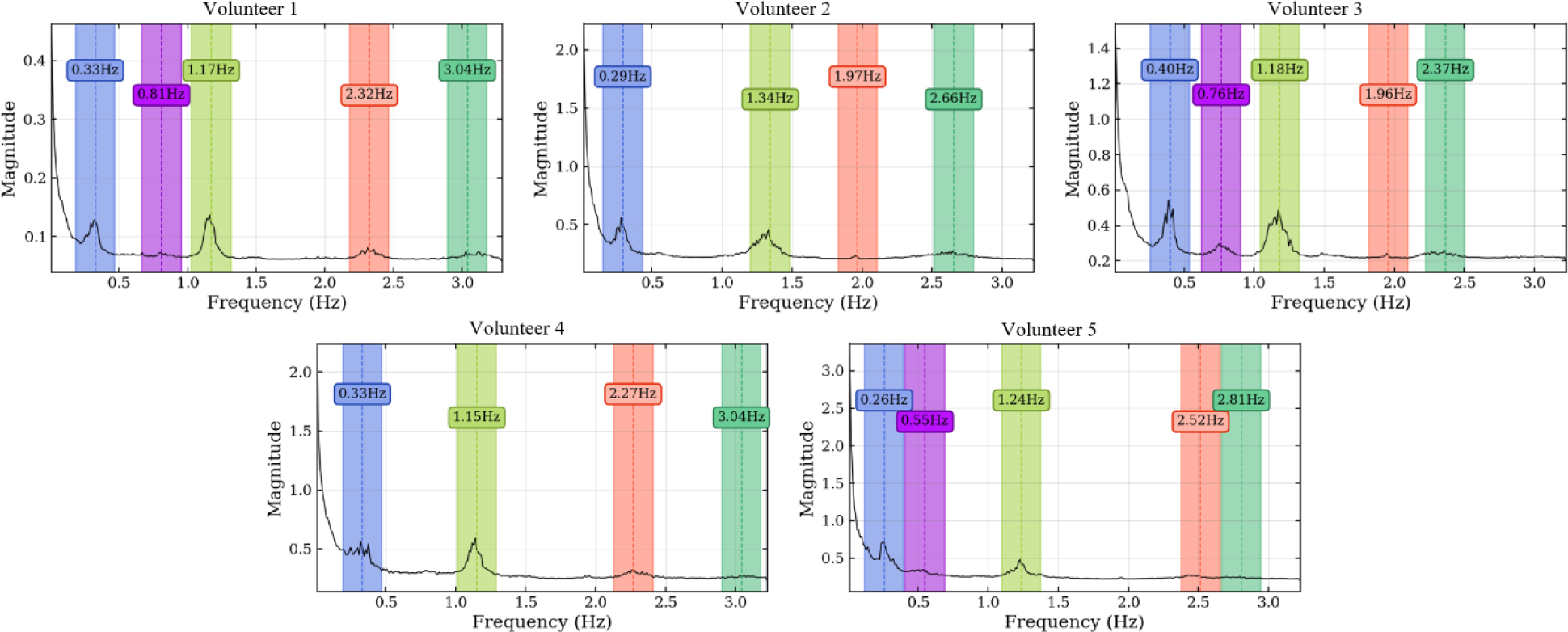
Frequency spectrum of the average signal intensity within the brain for 5 subjects. Regions of 0.3Hz around each peak frequency were highlighted to denote the bandwidth used for spatial mask creation.

**Table 1.** Values of the calculated center frequency responses for each identified frequency band.

|  | **Band 1** | **Band 2** | **Band 3** | **Band 4** | **Band 5** |
| --- | --- | --- | --- | --- | --- |
| **Volunteer 1** | 0.329 | 0.811 | 1.173 | 2.325 | 3.037 |
| **Volunteer 2** | 0.290 | - | 1.344 | 1.968 | 2.656 |
| **Volunteer 3** | 0.398 | 0.763 | 1.183 | 1.957 | 2.366 |
| **Volunteer 4** | 0.333 | - | 1.151 | 2.269 | 3.043 |
| **Volunteer 5** | 0.258 | 0.548 | 1.237 | 2.516 | 2.806 |

The masks created per frequency band (Figure 5) show a spatial localization for the frequency band centered at lower frequencies, e.g., 0.40Hz (Figure 5a) overlapping with brain regions with larger volume of CSF (the main ventricles and the brain periphery). Frequency bands centered at heart rate frequencies, e.g., 1.18Hz (Figure 5c) can be found on the regions with a stream of CSF (the main cerebral aqueduct). Similar patterns were observed for all volunteers as shown in Figure 6 as the mask for the heart rate band is demonstrated in each of the volunteer’s data.

**Figure 5:**
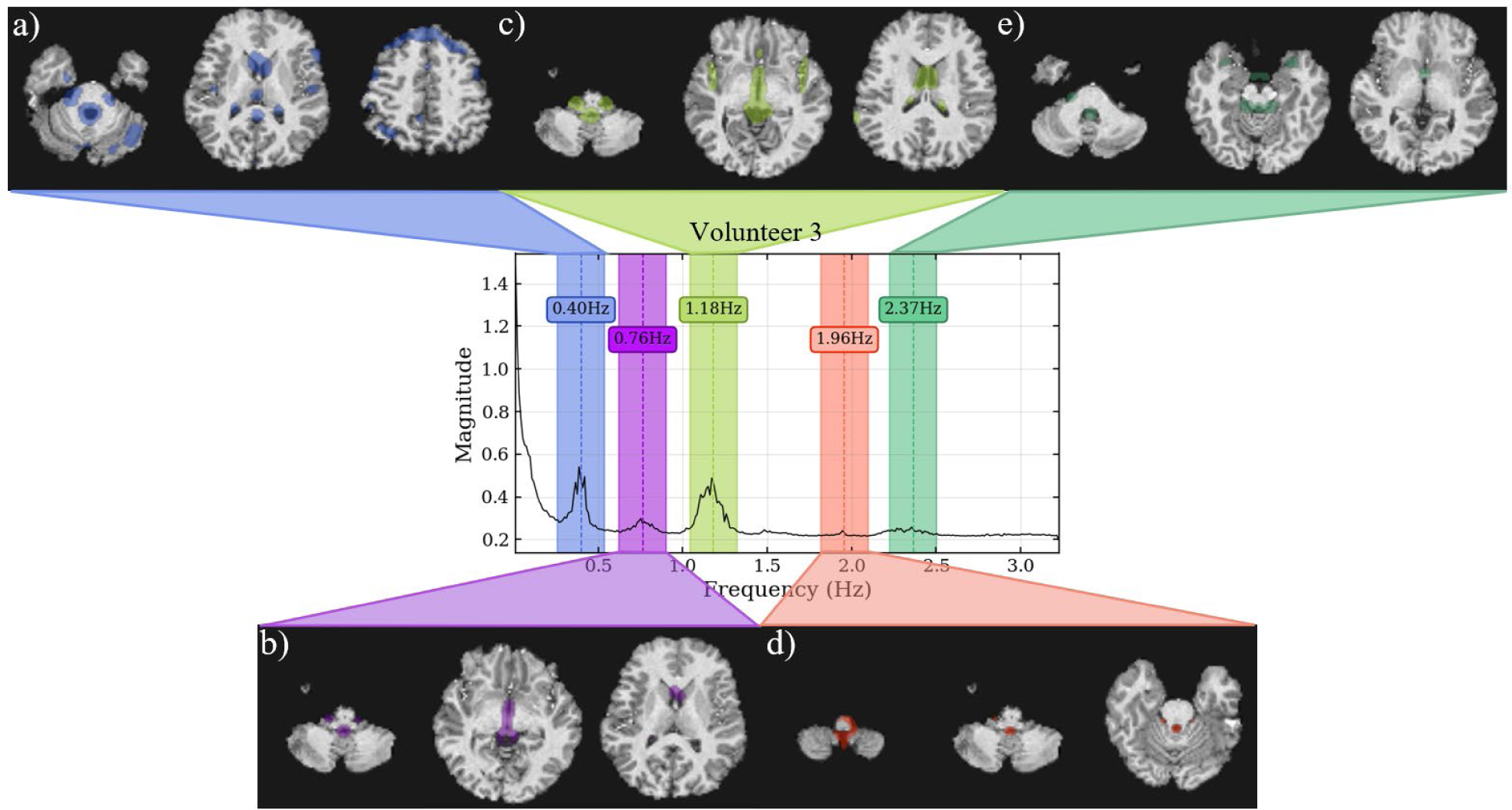
Frequency spectrum of volunteer 3 with spatial localization of the signal from 5 separate frequency bands. For each band, a spatial mask was applied to the T1 weighted image. The acquired data can be obtained from the inferior region of the brain (cerebellum) up to the middle of the brain. The bandwidth for each band is 0.3Hz. The center frequencies are a) 0.4Hz, b) 0.76Hz, c) 1.18Hz, d) 1.96Hz, and e) 2.37Hz. The acquisition was done using an EPI sequence with TR=155ms with 19 slabs of 3 slices each for a total of 57 slices.

**Figure 6:**
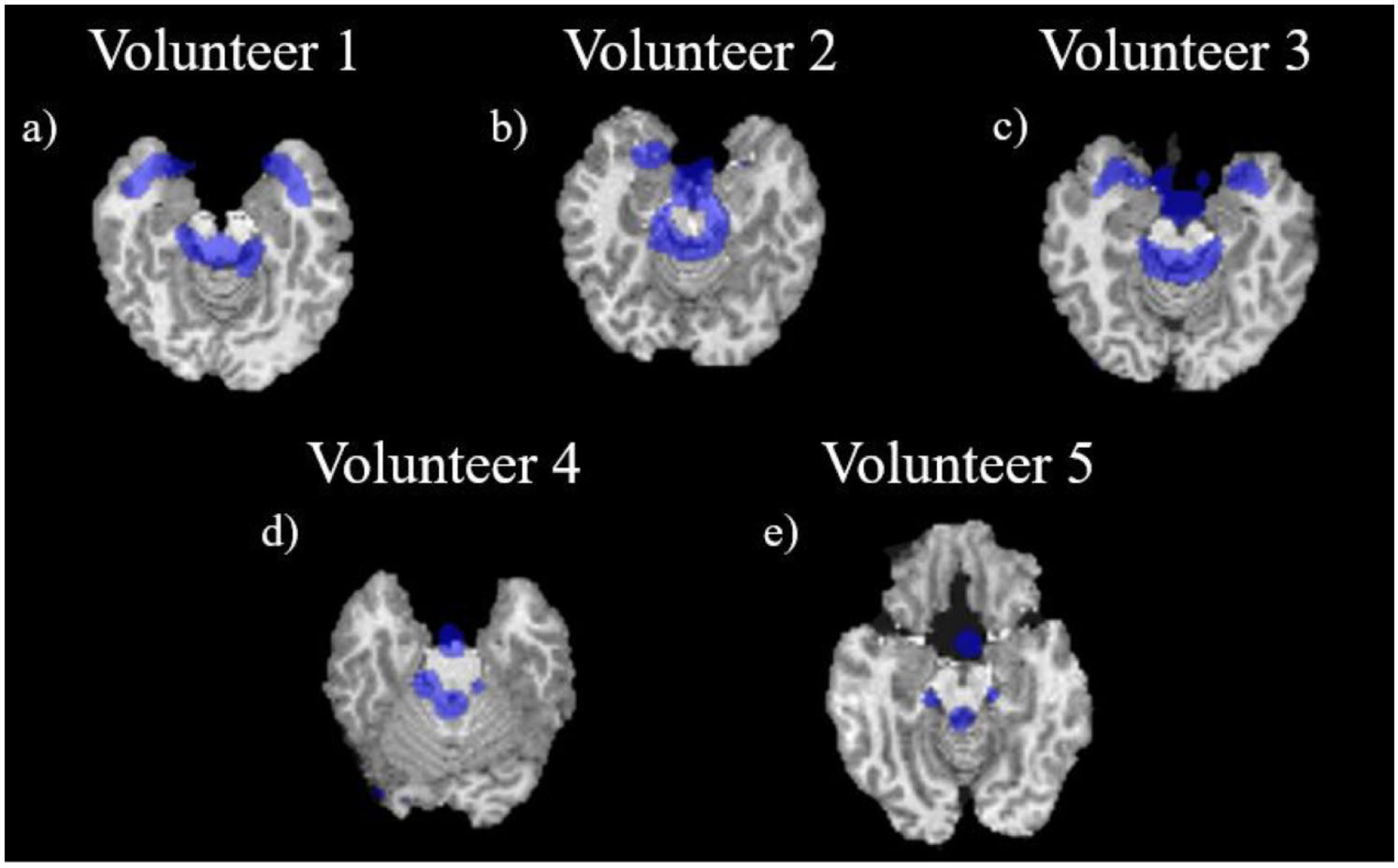
Visualization of the mask created for each volunteer at approximately the same position in the brain (bottom of the brain and top of the cerebellum and at approximately the same frequency band; a) for volunteer 1 at 1.17Hz; b) for volunteer 2 at 1.34Hz; c) for volunteer 3 at 1.18Hz; d) for volunteer 4 at 1.15Hz; and e) for volunteer 5 at 1.24Hz.

For the larger frequency spectrum (dataset with TR of 51ms), extra bands can be identified, and the center frequency of the most prominent band was calculated at 3.5Hz (Figure 7).

**Figure 7:**
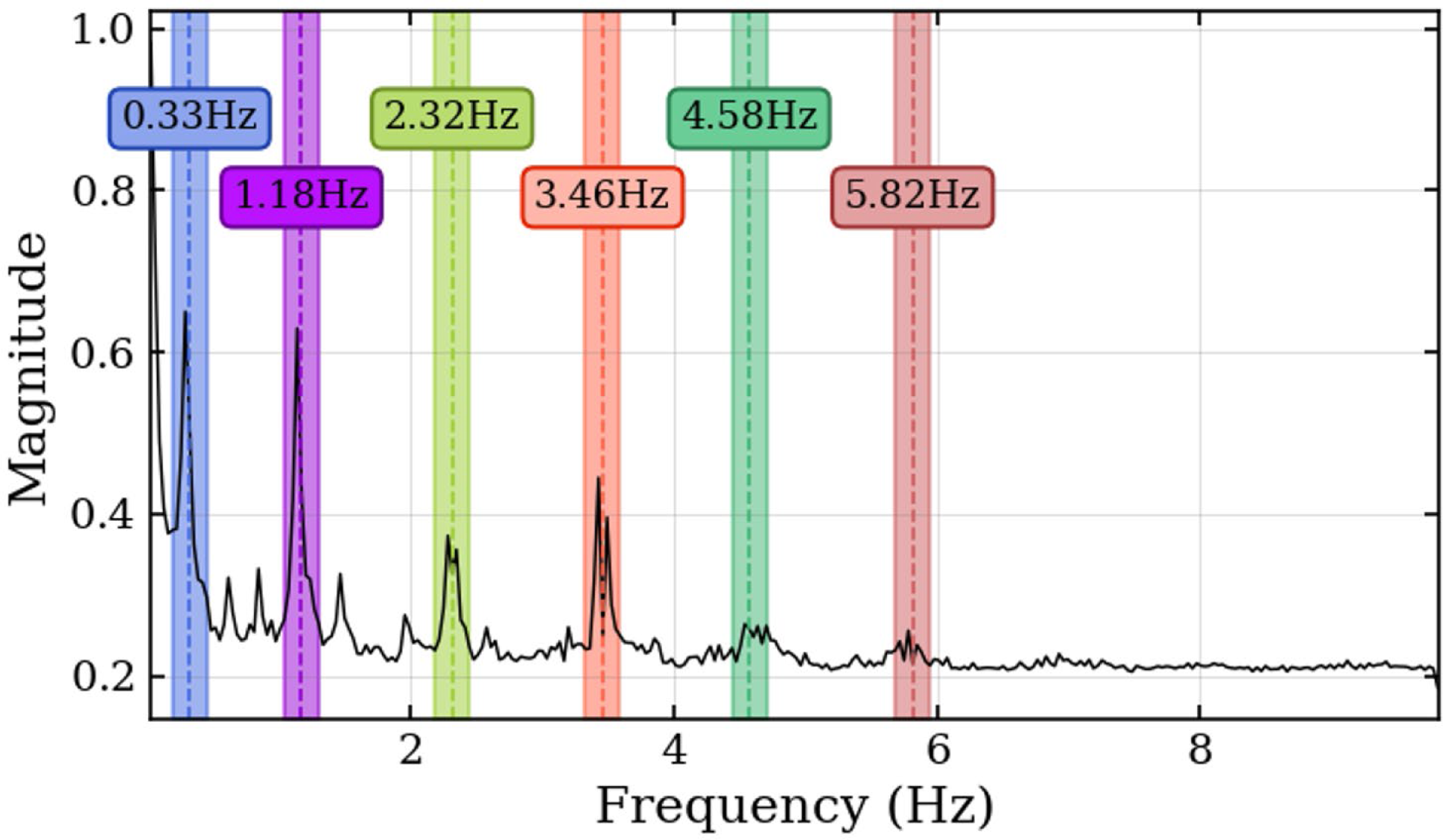
Frequency spectrum for volunteer 1 done using an EPI sequence single slice with TR=51ms. Maximum frequency of 9.8Hz and frequency bands of 0.3Hz were highlighted for better visualization of the main frequencies shown in the spectrum.

## 4. Discussion

We demonstrated a method to analyze the CSF motion in the human brain in-vivo using ultrafast 7T EPI acquisitions. The raw visualization of the real-time signal (Figure 1) shows *in-vivo* CSF motion. The flow of CSF within the ventricles and in the subarachnoid space can be clearly visualized with changes in signal intensity. The time series and the frequency spectrum comparison between the collected physiological data and the EPI data (Figure 2) shows a direct alignment between the two types of data where the cardiac and respiration cycles can be observed in the EPI MRI data. The frequency analysis also shows consistent results across multiple volunteers, with similar frequency spectrums are observed.

Compared to previous studies (Fultz et al., 2019; Shanks et al., 2019; Yamada et al., 2020; Yang et al., 2022), we were able to achieve a greater frequency range and improved frequency resolution. This was made possible by utilizing ultrafast acquisition times (ranging from 51ms to 150ms), which allowed for whole-brain spectral analysis up to 9.8Hz. Additionally, we employed high SNR and homogeneous images by using a 7T MRI with a customized RF coil system(Krishnamurthy et al., 2019; Santini et al., 2018; Santini, Wood, et al., 2021). To optimize sequence parameters, we tailored the flip-angle to maximize the signal of the CSF flow and adjusted other parameters to minimize susceptibility-related distortions. We also selected a TE that could potentially capture the BOLD signal if functional connectivity data are warranted. The creation of frequency masks allowed for an analysis of the spatial localization of each frequency band. The presence of lower frequencies responses (respiration band – 0.4Hz) in the brain periphery suggests a less turbulent fluid flow in regions with more space for fluid flow. Furthermore, the presence of heart rate frequencies (1.2Hz) in the ventricles further validates the analysis as the arterial pulse wave in the choroid plexus, for instance, is known to influence the CSF motion(Bilston et al., 2010; Iliff et al., 2013; Martin et al., 2012). Additionally, the presence of high frequencies (over 2Hz) responses can suggest a more turbulent flow that also aligns with regions of the main cerebral aqueduct.

This work provides a basis for potentially identifying new biomarkers for brain fluid dynamics. For example, the frequency spectrum can be analyzed for different brain diseases. The lower frequency bands (below 1Hz) contain physiological signals that corelate with the heart rate and breathing, so brain conditions that affect those variables can be analyzed directly from the MRI data. The magnitude of each band may also provide insights into the coupling between the heart and breathing rates with the CSF movement, which may indicate lower clearance rate. On the other hand, the higher frequency bands (above 1.8Hz), can be correlated with sleep cycles and potential sleep studies (Xie et al., 2013).

## 5. Conclusion

The development of non-invasive neuroimaging methods and biomarkers is essential for studying brain diseases. This work presented a novel methodology to characterize the frequency spectrum and spatial localization of CSF oscillations and movements in the human brain. The use of ultrafast EPI in conjunction with 7T human MRI and simultaneous collection of physiological data enabled the identification of primary components of CSF oscillations and their mapping spatially and temporally onto the MR image and physiological domains. The methodology showed good consistency and repeatability in-vivo, making it a promising tool for potential brain dynamics and CSF flow/clearance studies. These findings may have significant implications in the diagnosis and treatment of brain diseases, and further research is necessary to explore the potential of this methodology in clinical studies.

## Data and Code Availability

The codes and dataset used in this work are available upon request.

## Author Contributions

Conceptualization: TM, TS, MW, KAW, DM, JWI, HJA, TSI; Methodology: TM, TS, MW, JWI, TSI; Software: TM, TS, MW; Formal analysis: TM, TS; Investigation: TM, TS, JWI, TSI; Resources: JWI, TSI; Writing - Original Draft: TM, TS, KAW, DM, JWI, HJA, TSI; Writing - Review & Editing: TM, TS, KAW, DM, JWI, HJA, TSI; Supervision: TSI; Funding acquisition HJA, TSI.

## Data Availability

All data produced in the present study are available upon reasonable request to the authors

## Acknowledgments and Funding

This work was supported by NIH R01AG063525 and R01MH111265. The author Tiago Martins was partially supported by the CAPES Foundation, Ministry of Education of Brazil, 88881.128222/2016-01. This research was supported in part by the University of Pittsburgh Center for Research Computing, RRID:SCR_022735, through the resources provided. Specifically, this work used the H2P cluster, which is supported by NSF award number OAC-2117681.

